# Human milk short-chain fatty acid concentrations are not associated with early childhood allergic disease

**DOI:** 10.64898/2026.07.12.26357877

**Authors:** Lisa F. Stinson, Debra J. Palmer, Susan L. Prescott, Nina D’Vaz, Vaitheeswari, Kevin Huynh, Thy Duong, Peter J. Meikle, Donna T. Geddes, Alexandra D. George

## Abstract

**Background:** Short-chain fatty acids (SCFAs) are microbial metabolites with immunoregulatory properties. Human milk contains SCFAs which have been proposed as potential modulators of infant immune development. We aimed to examine associations between human milk SCFA concentrations and infant allergic disease outcomes in a high-risk cohort of infants of atopic mothers.

**Methods:** SCFAs were measured by targeted liquid chromatography-mass spectrometry in human milk samples collected at 3 and 6 months postpartum from atopic mothers enrolled in the Infant Fish Oil Supplementation (IFOS) Study (n=147). Associations between milk SCFA concentrations and early childhood allergic disease outcomes (atopic dermatitis, food allergy, allergic rhinitis, and allergen sensitisation at 1 and 2-3 years) were examined using logistic regression.

**Results:** Human milk SCFA concentrations were broadly stable between 3 and 6 months postpartum, except for acetate which was significantly elevated at 6 months. No significant associations were observed between human milk SCFA concentrations and any allergic disease outcome after correction for multiple comparisons (all p>0.05).

**Conclusions:** Human milk SCFA concentrations are not associated with allergic disease outcomes up to 4 years of age. These findings suggest that oral SCFA exposure via human milk is insufficient to reduce infant allergy risk, and that gut SCFA production may be a more relevant target for future allergy prevention research.

## Introduction

Infants born to atopic mothers face a substantially elevated risk of developing allergic disease in early childhood [1-3]. With allergic disease now affecting an estimated 30–40% of the population worldwide [4] and prevalence continuing to rise, identifying modifiable exposures that could reduce this risk is an urgent clinical and public health priority. The early postnatal period represents a critical window during which the immune system is shaped toward tolerance or allergy, making early intervention a promising avenue for prevention.

Among the modifiable exposures under investigation, short-chain fatty acids (SCFAs) have emerged as particularly promising candidates [5, 6]. SCFAs are small metabolites produced primarily through microbial fermentation of dietary fibre in the gut, that are then distributed systemically via the circulatory system. SCFAs have been shown to be potent regulators of immune development, with a growing body of preclinical and human evidence supporting their role in shaping allergic disease risk [6-14]. In murine models, maternal and early-life SCFA exposure has been shown to promote regulatory T cell (Treg) development, suppress type 2 inflammatory responses, and reduce allergen sensitisation and airway inflammation [7, 13, 14]. *In-vitro* studies have demonstrated that butyrate and propionate modulate dendritic cell and epithelial cell function, enhancing barrier integrity and dampening Th2 cytokine production [14-17]. In human cohort studies, higher circulating SCFA concentrations in infancy have been associated with reduced risk of allergic sensitisation, food allergy, and atopic eczema in early childhood [12], and gut SCFA production has been inversely associated with atopic disease development [6, 9-11]. Collectively, this evidence positions SCFAs as biologically plausible candidates for early immune programming to reduce the risk of allergic disease development.

Human milk contains measurable SCFAs [8, 12, 18, 19], and has therefore been proposed as a potential mediator infant immune protection [5]. Critically, if human milk derived SCFAs can shape infant immune development, maternal dietary fibre supplementation during lactation could represent a simple, low-risk intervention to increase human milk SCFA and reduce allergy risk in susceptible infants. To date, limited research has directly examined associations between human milk SCFA concentrations and infant allergy outcomes. One small case-control study (n=47 per group) reported that acetate concentrations in human milk were negatively associated with atopic dermatitis in infancy [8]. However, a larger prospective cohort (n=148) found no association between milk SCFAs and allergic outcomes at 12 months, despite protective associations for infant plasma SCFAs [12]. Taken together, the available evidence remains limited and inconsistent, highlighting the need for larger prospective studies to determine whether milk-derived SCFAs represent a biologically relevant exposure for infant immune development. Here, we aimed to examine associations between human milk SCFA concentrations at 3 months postpartum and infant allergy outcomes at 1 and 2-3 years in a high-risk cohort of infants of allergic mothers.

## Methods

### Study Population

This study utilised human milk samples and clinical data from the Infant Fish Oil Supplementation (IFOS) Study, a double-blind randomised controlled trial conducted in Perth, Western Australia, designed to assess the effects of infant fish oil supplementation on allergy and neurodevelopmental outcomes (ACTRN12606000281594). The study design and methodology have been described in detail elsewhere [20]. Ethical approval was obtained from the Princess Margaret Hospital Human Research Ethics Committee (Approval #1111EP), and all participants provided written informed consent. Briefly, atopic mothers were recruited at 36 weeks of pregnancy from metropolitan antenatal clinics in Perth between 2005 and 2008. Eligible mothers were defined as atopic by a positive skin prick test to at least one common allergen and a history of doctor-diagnosed allergic disease (food allergy, asthma, allergic rhinitis, or atopic dermatitis). The inclusion of atopic mothers enriches the population for allergic outcomes, enhancing statistical power to detect associations where they exist. As the IFOS intervention was administered to infants and did not influence infant allergic outcomes [21], participants from both the fish oil and control groups were included in the present analysis. Of the mothers enrolled in the IFOS Study, 281 had human milk samples available for analysis. A subset of samples (n = 67) was excluded due to a cold chain breach during storage, which was considered likely to affect SCFA composition. The final sample set for this study comprised 148 participants (147 with samples at 3 months, 147 at 6 months, and 146 with samples at both time points) (**Supplementary Figure 1**). Maternal and infant characteristics of the analytic sample set are summarised in **Supplementary Table 1**.

### Human Milk Sample Collection

Human milk samples were collected at 3 and 6 months postpartum by either hand expression or breast pump. Mothers were instructed to collect up to 6 mL of human milk prior to a morning feed. Samples were immediately frozen and stored at -80°C, undergoing a single freeze-thaw cycle prior to analysis.

### Allergic Disease Outcomes Assessment

Child allergic disease outcomes were assessed by clinical examination at 1 year and 2-3 years of age. Outcomes included doctor-diagnosed atopic dermatitis, food allergy, and allergic rhinitis. Allergic sensitisation was assessed by skin prick test to a panel of common food and environmental allergens at 1 year and 2-3 years, with sensitisation defined as a wheal diameter of ≥2 mm to at least one allergen. Additionally, a composite variable for “any allergic disease” was derived for each time point, defined as positive at 1 year if the participant had either atopic dermatitis or food allergy, and as positive at 2 years if the participant had atopic dermatitis, food allergy, or allergic rhinitis. For each outcome, a composite “ever” variable was derived, defined as a positive outcome at either 1 year or 2-3 years of age. Participants were classified as positive if they met the outcome criterion at either time point (irrespective of whether data were available at the other time point), negative only if they were negative at both time points and missing if one time point was negative and the other was unavailable.

### Short Chain Fatty Acid analysis

Human milk samples were thawed at room temperature and mixed thoroughly prior to extraction. The following methodology was developed from similar published methods [22-24]. A 10 µL aliquot of human milk was combined with 10 µL of internal standard mix and diluted with 50 µL of 75% methanol. The derivatising reagents (10 µL of each, 200 mM 3-NPH and 120 mM EDC in 6% pyridine) were added and samples were kept at room temperature for 45 min with shaking. The reaction was quenched with 50 µL of 200 mM quinic acid, 15 minutes shaking, then centrifuged at 15,000 x *g* for 5 min. The supernatant (90 µL) was transferred to a glass vial and diluted to a final volume of 1 mL with 10% methanol in water (v/v) for analysis. SCFAs were quantified using targeted liquid chromatography–tandem mass spectrometry (LC-MS/MS) (Agilent 1290 UHPLC with an Agilent 6495C triple quadrupole mass spectrometer). Chromatographic separation was performed on a ZORBAX eclipse plus C18 column (2.1 x 100 mm, 1.8 µm, Agilent) with a binary solvent system of solvent A (water with 0.1% formic acid) and solvent B (95% acetonitrile with water + 0.1% formic acid). The gradient (0.8 mL/min) started with 95% solvent A, decreased to 90% over 1 minute, to 75% up to 5 minutes, then to 30% up to 10 minutes, followed by return to 95% for re-equilibration. Analytes were monitored using predefined precursor-to-product ion transitions specific to derivatised SCFAs, in positive ion mode, with the characteristic product ion, *m/z* 137.1, corresponding to the 3-NPH moiety [25]. The panel included SCFAs ranging from formate to caproate, including branched-chain isomers, alongside their corresponding isotopically labelled internal standards. For the purposes of this study, formate was included as a retention time place holder and caproate was considered a SCFA. Analytical performance was assessed using technical (CV% < 6.5%) and pooled batch (CV% < 15%) quality control samples distributed throughout the run. All SCFA were confirmed linear up to 1000 µM (R^2^ ≥ 0.94). Blank injections were included every 20 samples to monitor contamination and carryover. For each SCFA, chromatographic peaks were integrated using Mass Hunter (B.09.00, Agilent Technologies) software. Concentrations were calculated based on area under the chromatographic curve relative to the labelled internal standard concentrations and median blank concentrations were subtracted from each sample.

### Statistics

All statistical analyses were performed in RStudio (v 4.5.1) [26]. SCFA concentrations were log-transformed prior to all analyses to account for right-skewed distributions.

To examine whether human milk SCFA concentrations differed between 3 and 6 months postpartum, paired comparisons were performed for each SCFA using the subset of participants with samples at both time points (n=146). Prior to testing, the normality of pairwise differences (6M - 3M) was assessed using the Shapiro-Wilk test. Where differences were normally distributed (p>0.05), a paired t-test was applied; otherwise a Wilcoxon signed-rank test was used. P-values were adjusted for multiple comparisons using the Benjamini-Hochberg (BH) method.

Although SCFA concentrations were measured at both 3 and 6 months, regression analyses were restricted to the 3-month time point. SCFA concentrations were broadly stable across this period, with only acetate differing significantly between time points (see Results), suggesting that 3-month measurements are representative of the early lactation SCFA profile. The 3-month time point was additionally preferred as exclusive breastfeeding rates were higher, minimising dietary confounding from mixed feeding and introduction of solid food.

Potential confounders were screened for association with human milk SCFA concentrations at 3 months prior to model building. Variables assessed included birth season, delivery mode, infant sex, parity, and feeding mode at 3 months. Categorical variables were tested against log-transformed SCFA concentrations using the Kruskal-Wallis test, and parity was assessed using Spearman correlation. Multiple testing correction was applied using the BH method [27]. As samples were run in two batches, all models were adjusted for analytical batch.

Associations between human milk SCFA concentrations at 3 months and child allergic disease outcomes were examined using logistic regression fitted using the glm function (base R stats package) with a binomial family and logit link. Separate models were fitted for each SCFA-outcome combination, with each SCFA entered as a continuous exposure after log-transformation and z-score standardisation, allowing odds ratios to be interpreted per standard deviation increase in SCFA concentration. All models were adjusted for analytical batch. For each analysis, cases with missing data were excluded. To account for multiple testing across SCFA-outcome combinations, BH correction was applied separately within each outcome. Results are reported as odds ratios (OR) with 95% confidence intervals (CI) with both unadjusted and BH-corrected p values. Sensitivity analyses were conducted to assess the robustness of findings to feeding mode. Specifically, models were additionally adjusted for feeding mode at 3 months and age at cessation of breastfeeding, and results were compared to the primary models.

## Results

A total of 147 mother-infant pairs with human milk available at 3 months postpartum were included in the analysis. The prevalence of allergic disease outcomes at 1 and 2-3 years of age is summarised in **Table 1**. In this high-risk cohort, over half (52.4%) of the infants experienced an allergic manifestation within the first two years of life, with the most common being atopic dermatitis (44.2%).

**Table 1.** Prevalence of allergic disease outcomes at 1 and 2-3 years of age (n = 147). Data are presented as n (%).

| Infant outcome | No | Yes | Missing |
| --- | --- | --- | --- |
| Sensitisation (1Y) | 98 (66.7%) | 42 (28.6%) | 7 (4.8%) |
| Sensitisation (2-3Y) | 82 (55.8%) | 36 (24.5%) | 29 (19.7%) |
| Sensitisation (ever) | 68 (46.3%) | 55 (37.4%) | 24 (16.3%) |
| Any allergic disease (1Y) | 54 (36.7%) | 63 (42.9%) | 30 (20.4%) |
| Any allergic disease (2-3Y) | 53 (36.1%) | 64 (43.5%) | 30 (20.4%) |
| Any allergic disease (ever) | 30 (20.4%) | 85 (57.8%) | 32 (21.8%) |
| Food allergy (1Y) | 89 (60.5%) | 27 (18.4%) | 31 (21.1%) |
| Food allergy (2-3Y) | 91 (61.9%) | 27 (18.4%) | 29 (19.7%) |
| Food allergy (ever) | 67 (45.6%) | 39 (26.5%) | 41 (27.9%) |
| Atopic dermatitis (1Y) | 68 (46.3%) | 54 (36.7%) | 25 (17.0%) |
| Atopic dermatitis (2-3Y) | 75 (51.0%) | 42 (28.6%) | 30 (20.4%) |
| Atopic dermatitis (ever) | 48 (32.7%) | 65 (44.2%) | 34 (23.1%) |
| Allergic rhinitis (2-3Y) | 91 (61.9%) | 27 (18.4%) | 29 (19.7%) |

Human milk SCFAs exhibited high variation across all samples, ranging from lowest, 0.16 µmol/L (valerate), to highest, 2259.52 µmol/L (caproate) (**Table 2**). Human milk SCFA concentrations were largely stable across the 3 and 6 month samples (**Figure 1**). Of the seven SCFAs examined, only acetate differed significantly over time, increasing from 3 to 6 months postpartum with a moderate effect size (r=0.395, BH-adjusted p=0.00001).

**Figure 1:**
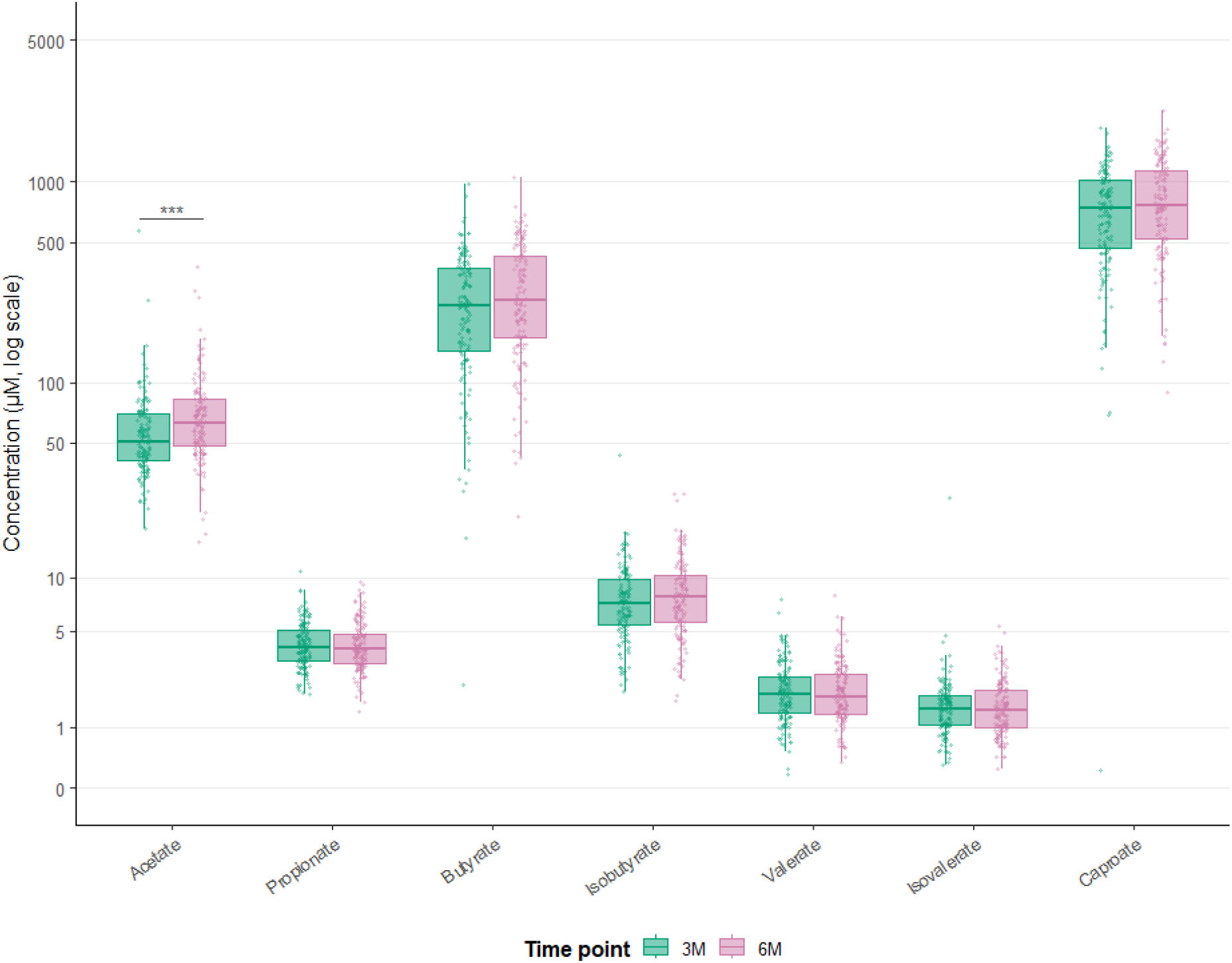
Human milk short-chain fatty acid concentrations at 3 and 6 months postpartum. Concentrations (µM) of SCFAs measured by LC-MS in Human milk collected at 3 months (teal) and 6 months (pink) postpartum. *** BH-adjusted p>0.001.

**Table 2.** Descriptive statistics for human milk SCFA concentrations (3 and 6 months).

| SCFA species | Mean ± SD (range) ($\mu\text{mol/L}$) |
| --- | --- |
| Acetate | 67.3 ± 49.0 (15.5-572.9) |
| Propionate | 4.2 ± 1.5 (1.4-10.9) |
| Butyrate | 285.4 ± 173.0 (2.3-1048.0) |
| Isobutyrate | 8.2 ± 4.4 (1.7-43.1) |
| Valerate | 2.1 ± 1.1 (0.2-8.0) |
| Isovalerate | 1.6 ± 1.6 (0.2-26.3) |
| Caproate | 796.3 ± 390.9 (0.2-2259.5) |

No significant associations were observed between human milk SCFA concentrations at 3 months and any allergic disease outcome at 1 or 2-3 years after correction for multiple comparisons (**Figure 2**). Prior to correction, acetate was positively associated with food allergy at 1 year (OR 1.69 [95% CI 1.06–2.69], p=0.028), 2-3 years (OR 1.67 [95% CI 1.01–2.77], p=0.046), and ever (OR 1.69 [95% CI 1.01–2.81], p=0.044), as well as any allergy at 1 year (OR 1.59 [95% CI 1.03–2.47], p=0.037) and ever (OR 1.79 [95% CI 1.02-3.14], p=0.042). However, no finding remained after BH correction (all BH-adjusted p>0.15). Full results for all SCFA-outcome combinations are presented in **Supplementary Table 2**.

**Figure 2:**
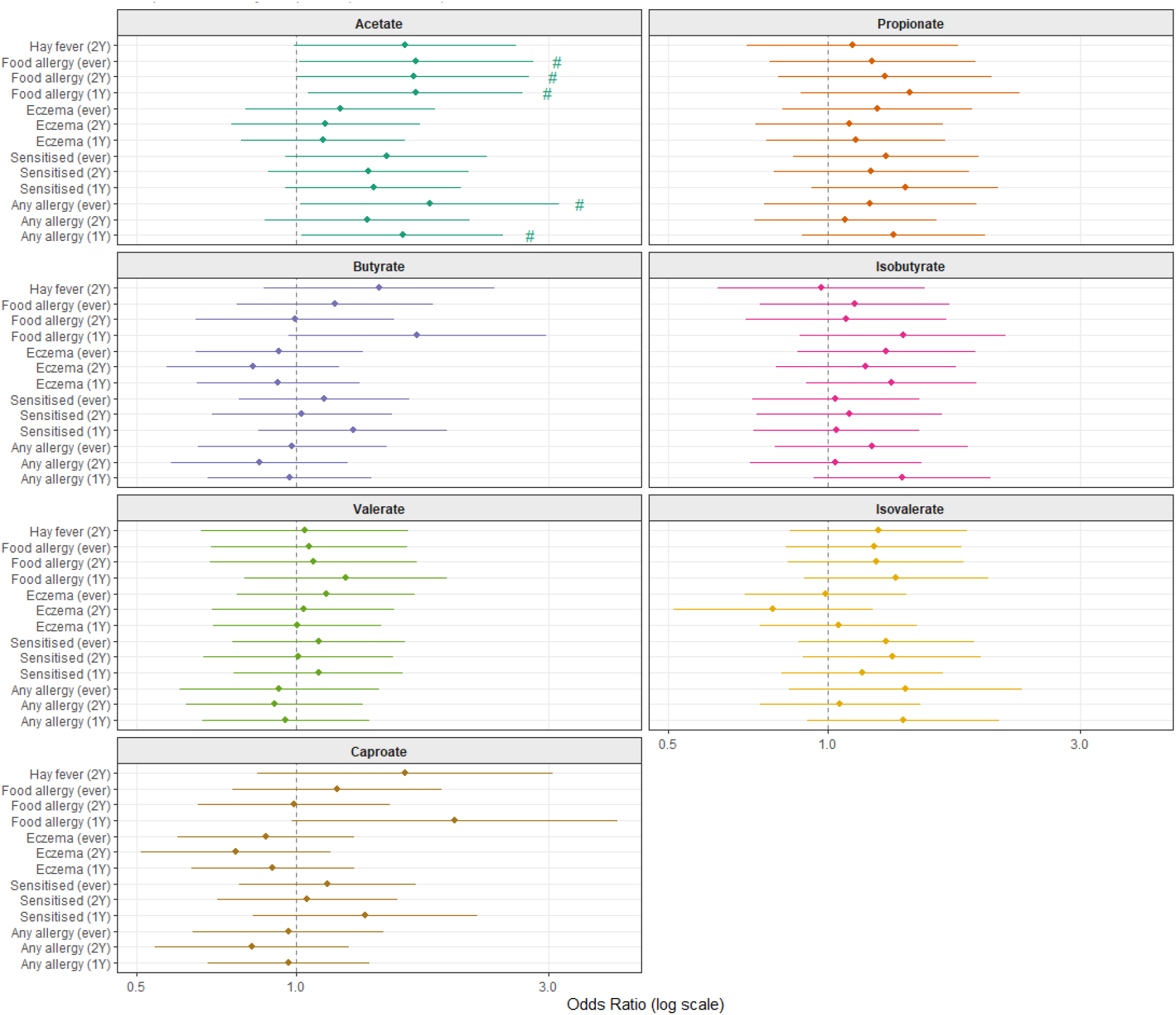
Associations between human milk short-chain fatty acid concentrations at 3 months postpartum and infant allergic disease outcomes at 1 and 2-3 years. Odds ratios (95% CI) are derived from logistic regression models. # unadjusted p<0.05.

Sensitivity analyses adjusting for feeding mode at 3 months and total duration of breastfeeding did not alter these findings, and no associations reached statistical significance after BH correction. Results of sensitivity analyses are presented in **Supplementary Tables 3 and 4**.

## Discussion

Human milk SCFAs have been postulated to have a role in modulating allergy risk in early life. In this prospective cohort of infants born to atopic mothers, we found no evidence that human milk SCFA concentrations at 3 months postpartum were associated with child allergic disease outcomes at 1 or 2-3 years of age. These findings are consistent with those of Barman *et al*., who similarly reported no association between milk SCFA concentrations and infant allergic sensitisation or atopic disease outcomes in a prospective cohort, despite observing associations between infant plasma SCFAs and these same outcomes [12]. In contrast, a prior case–control study reported associations between milk acetate and atopic dermatitis; however, this study was limited by small sample size and retrospective design, reducing its ability to infer temporality [8]. Together, the available prospective evidence converges on a null relationship between human milk-derived SCFAs and infant allergy outcomes.

These results raise an important mechanistic question: if SCFAs are immunoprotective in early life, why does their presence in human milk not confer benefit? We propose that two factors, the route of delivery and the dose of exposure, may together explain why milk-derived SCFAs are insufficient to exert meaningful immunomodulatory effects. With respect to route, milk-derived SCFAs represent an exogenous oral exposure rather than endogenous colonic production. Unlike gut-derived SCFAs, which are produced in the colon and exert local and systemic immune effects via direct interaction with colonic epithelium, dendritic cells, and regulatory T cells, milk-derived SCFAs represent an exogenous oral exposure rather than endogenous microbial production. This distinction may be biologically important, as milk-derived SCFAs are delivered orally and are likely absorbed in the proximal gastrointestinal tract before reaching the colon [28]. This is consistent with pharmacokinetic evidence in adults demonstrating that orally consumed SCFAs reach peak plasma concentrations within 60 minutes, suggesting rapid proximal absorption [28]. As such, clinical trials that involve oral SCFA supplementation conjugate them to high-amylose maize starch to ensure they reach the colon [29, 30]. If milk-derived SCFAs are similarly absorbed in the proximal gut of the breastfed infant, they may be metabolised before reaching the colon. Indeed, Barman *et al*. found that milk SCFA concentrations did not correlate with infant plasma SCFA concentrations, suggesting that milk-derived SCFAs make little contribution to the infant’s systemic SCFA pool [12]. With respect to dose, the quantities of SCFAs delivered via human milk are likely insufficient to drive immunological effects even if they were to reach the colon. The immunological effects of SCFAs demonstrated in preclinical models have predominantly been observed at concentrations far exceeding those present in human milk. Butyrate concentrations used in murine models of food allergy are estimated to exceed infant exposure via human milk by several hundred-fold, questioning the translational relevance of these findings to breastfed infants. Furthermore, the absolute quantity of SCFAs delivered via human milk is likely small relative to endogenous production in the infant gut. Assuming an infant consumes 750 mL/day of milk, this corresponds to a total SCFA intake of approximately 0.9 mmol/day. This can be compared to gut SCFA production of 5-10 mmol/day. This estimate suggests that milk-derived SCFAs represent only a minor fraction (approximately 5–10%) of total SCFA exposure, with the majority generated through microbial fermentation in the colon [31, 32]. Collectively, both the route and quantity of SCFA delivery via human milk suggest that this source is unlikely to replicate the immunomodulatory effects associated with colonic SCFA production.

Consistent with these findings, randomised controlled trials targeting maternal SCFA production through prebiotic supplementation have reported null effects on infant allergic outcomes, despite successfully increasing maternal SCFA levels. In the SYMBA trial, maternal supplementation with galacto-oligosaccharides and fructo-oligosaccharides (GOS/FOS) from mid-pregnancy to 6 months postpartum did not reduce infant eczema [33], despite increases in maternal fecal SCFAs [34]. Similarly, in the PREGRALL trial, maternal supplementation with GOS/inulin from mid-pregnancy until delivery failed to prevent atopic dermatitis at 1 year in infants of atopic mothers [35]. Taken together with the observational evidence presented here and in Barman *et al*. [12], these findings suggest that maternal dietary fibre supplementation does not meaningfully influence infant allergy risk through modification of milk SCFA composition, and that this pathway is unlikely to represent an effective target for allergy prevention. In contrast, SCFAs produced within the infant gut remain a plausible and biologically relevant target. Associations between infant plasma or stool SCFA concentrations and allergic outcomes have been consistently reported [6, 9-12]. This distinction suggests that endogenous microbial production, rather than exogenous delivery via human milk, is the primary pathway through which SCFAs may influence immune development. Importantly, human milk may still contribute to this process indirectly. Human milk oligosaccharides (HMOs) serve as key substrates for *Bifidobacterium*-mediated fermentation in the infant colon, driving the production of SCFAs and other immunomodulatory metabolites during a critical window of immune development. In this light, the present findings do not negate a role for SCFAs in allergy prevention but rather refine the locus of interest from milk SCFA concentration to infant gut SCFA production. Future research should prioritise interventions that enhance endogenous SCFA production within the infant gut, such as synbiotic supplementation targeting SCFA-producing bacteria, or strategies that optimise HMO utilisation, where there is stronger mechanistic and epidemiological support for a protective effect.

The SCFA concentrations measured in this study were, on average, comparable to those previously reported in human milk studies, however very wide ranges have been described here and in other works [12, 14, 19, 36, 37]. Our analytical method incorporated several quality control measures, including the use of internal standards for each species, blank subtraction, low technical variation (mean CV 6.5%), and assessment of linearity and reproducibility, supporting the reliability of these measurements. However, SCFA quantification in human milk is sensitive to many pre-analytical factors, including sample collection, storage duration, freeze–thaw cycles, and potential microbial activity. These factors may contribute to variability across studies. For example, processing conditions may influence the measured free SCFA pool, through hydrolysis of esterified lipids.

Our study, which is the largest human cohort to examine milk SCFAs and infant outcomes to date, has several strengths. These include its prospective design, the focus on a high-risk cohort born to atopic mothers, outcomes measured across two points, and the comprehensive quantification of multiple SCFAs using a validated sensitive LC-MS approach. However, several limitations should be considered. SCFAs are susceptible to variability related to sample handling and storage. In addition, we did not measure infant gut or plasma SCFA concentrations, limiting our ability to directly compare exogenous and endogenous SCFA exposure. Residual confounding is also possible. Finally, the actual intake of SCFAs by the infant was not quantified, as milk intake was not measured.

In conclusion, human milk SCFA concentrations were not associated with child allergic disease outcomes in this high-risk cohort. These findings, together with evidence from prospective studies and maternal prebiotic intervention trials, suggest that milk-derived SCFAs are unlikely to play a major role in allergy prevention. Future research should prioritise strategies that enhance endogenous SCFA production within the infant gut, such as targeted synbiotic supplementation or optimisation of HMO utilisation, rather than modification of maternal diet or milk composition.

## Data Availability

All data produced in the present study are available upon reasonable request to the authors

