## Supplementary Figure 1 for "Human milk short-chain fatty acid concentrations are not associated with early childhood allergic disease"

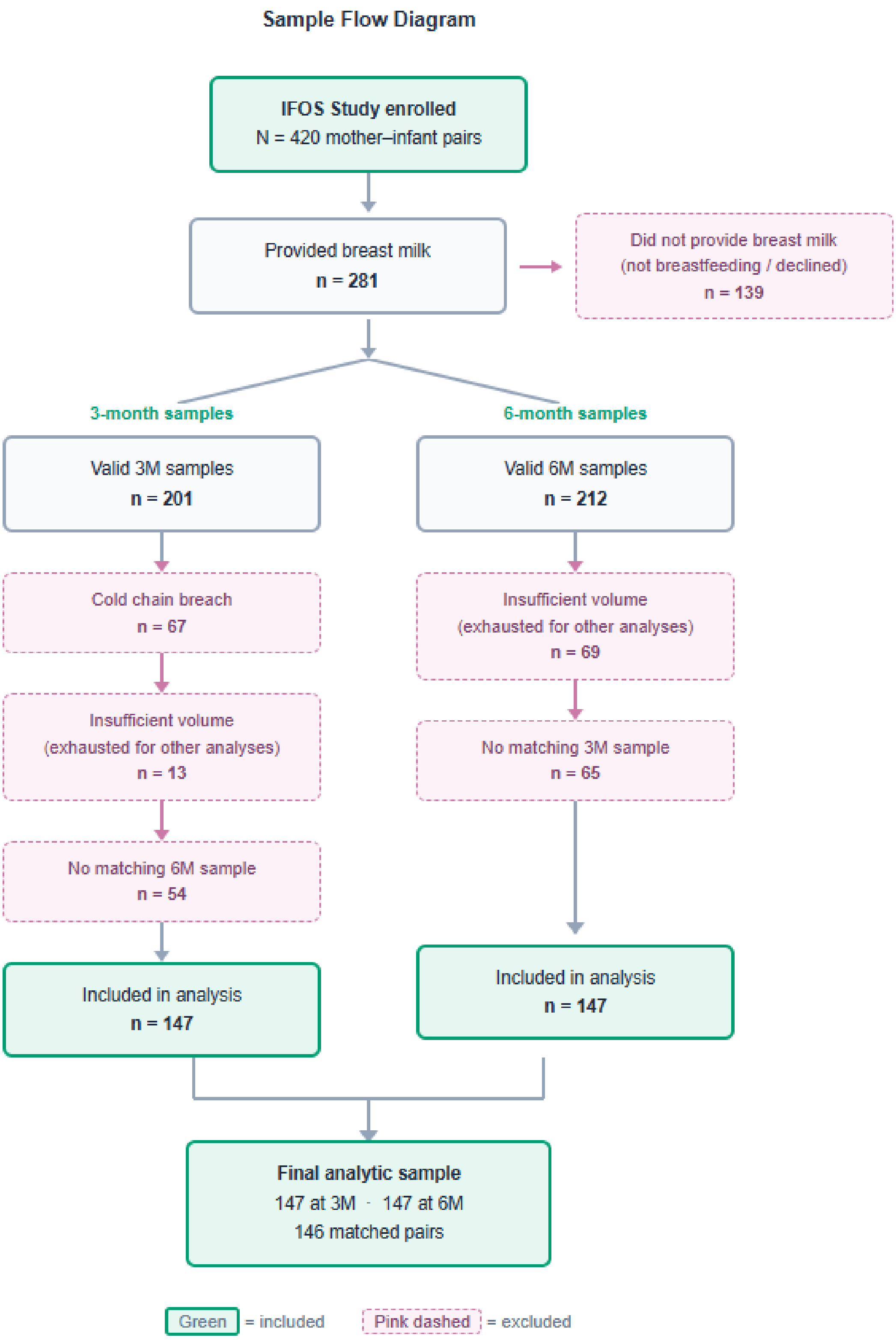


**Supplementary Figure 1:** Flow diagram of sample selection from the Infant Fish Oil Supplementation (IFOS) Study.
